# Lockdowns to Contain COVID-19 Increase Risk and Severity of Mosquito-Borne Disease Outbreaks

**DOI:** 10.1101/2020.04.11.20061143

**Authors:** Akshay Jindal, Shrisha Rao

**Affiliations:** Department of Computer Science and Technology, University of Cambridge, Cambridge CB3 0FD, UK.; International Institute of Information Technology - Bangalore, Bangalore 560 100, India.

## Abstract

Many countries are implementing lockdown measures to slow the COVID-19 pandemic, putting more than a third of the world’s population under restrictions. The scale of such lockdowns is unprecedented, and while some effects of lockdowns are readily apparent, it is less clear what effects they may have on outbreaks of serious communicable diseases. We examine the impact of these lockdowns on outbreaks of mosquito-borne diseases. Using an agent-based model and simulations, we find that the risk and severity of such outbreaks is much greater under lockdown conditions, with the number of infected people doubling in some cases. This increase in number of cases varies by different mosquito-borne diseases, and is significantly higher for diseases spread by day-biting mosquitoes. We analysed various intervention strategies and found that during lockdowns, decentralised strategies such as insecticide-treated nets and indoor residual spraying are more effective than centralised strategies.

## 1 Introduction

The ongoing COVID-19 pandemic caused by the coronavirus SARS-CoV-2 has led to large-scale lockdowns and quarantines all over the world, as authorities take recourse to such unprecedented measures to contain the spread of the disease. While some effects of these lockdowns are readily apparent— in the disruptions in economic activity, travel, education, and sports—and even in the undesirable consequences for the medical care of people with chronic or non-COVID illnesses, it is less obvious what immediate effects such lockdowns may have on outbreaks of other serious diseases.

Mosquito-borne diseases such as malaria, dengue fever, yellow fever, zika, chikungunya, and Japanese encephalitis are well known to afflict large parts of the world, and to extract a terrible toll in lives lost.

Using an agent-based model of mosquito-borne diseases that we have previously reported [9], we analyzed the effects of social lockdowns on the outbreaks of mosquito-borne diseases, and found that such lockdowns significantly increase the risk and severity of outbreaks of such diseases. The extent of such increases vary across diseases and lockdown configurations, but overall lockdowns are consistently worse for such diseases than the no-lockdown situation.

We also analyzed different intervention strategies to mitigate such mosquito-borne disease outbreaks during lockdowns, and found that unlike in a no-lockdown situation, decentralized strategies such as insecticide-treated nets (ITNs) and indoor residual spraying (IRS) are more effective than centralized strategies.

## 2 Results

To understand the implications of lockdowns for mosquito-borne diseases, we first investigate the impact of lockdowns on the risk and severity of outbreaks of such diseases. We may describe *risk* as the probability of starting an epidemic [7] upon the introduction of infection into the population through a single infected human agent. *Severity* is the total fraction of the local populace that are infected during the course of an outbreak.

We studied these impacts by running simulations of three of the most prevalent mosquito-borne diseases: chikungunya, dengue fever, and malaria. We ran agent-based simulations for each disease on a dataset describing St. Barth é emy (a small tropical island in the Caribbean), similar to our previous work [9]. However, we ran the model for three types of lockdown: (i) a full lockdown where the mobility of all human agents in the simulation is restricted to their homes; (ii) a partial lockdown (90%) where only 10% of the working human population (who may be essential workers) are allowed to carry out their day-to-day activities; and (iii) another partial lockdown (95%) where the same applies to only 5% of the working human population.

We report the findings below.

### There is greater risk and severity of outbreaks in lockdowns

Table 1 reports the increase in risk and severity for three lockdown configurations relative to the no-lockdown configuration. We note that both the risk and severity of outbreaks increases in all lockdown scenarios. This increase is significantly higher for diseases whose vectors are day-biting mosquitoes, i.e., chikungunya and dengue.

**Table 1:** Increase in the total number of cases (severity) and the probability of the infection spreading from the first carrier to the rest of the population(Risk), compared to the no-lockdown configuration

| Disease | Simulation Configuration |  |  |  |  |  |
| --- | --- | --- | --- | --- | --- | --- |
|  | Full Lockdown |  | Partial Lockdown (90%) |  | Partial Lockdown (95%) |  |
|  | Increase in Risk | Increase in Severity | Increase in Risk | Increase in Severity | Increase in Risk | Increase in Severity |
| Chikungunya | 37.5% | 62.5% | 25% | 162.5% | 25% | 87.5% |
| Dengue | 33.3% | 40% | 33.3% | 100% | 33.3% | 50% |
| Malaria | 25% | 18.2% | 25% | 45.5% | 25% | 36.4% |

### Partial lockdowns are worse than full lockdown

As seen in Table 1, the strictness of the lockdown also has a clear effect on the results. Partial lockdowns carry similar risks as the full lockdown but are much more severe.

### Trajectories for different diseases vary

We summarize the varying trajectories for different diseases as follows.

#### Chikungunya

In Figure 1, we plot the growth of epidemics for each disease and lockdown configuration. For chikungunya (Figure 1a), we see that the initial growth rate of an outbreak in lockdowns is significantly steeper than with no lockdown. Outbreaks in lockdown are also more localized in nature with a higher chance of recurrence, as seen by plateaus followed by a steep rise in the plots. This effect is enhanced in partial lockdowns.

**Figure 1:**
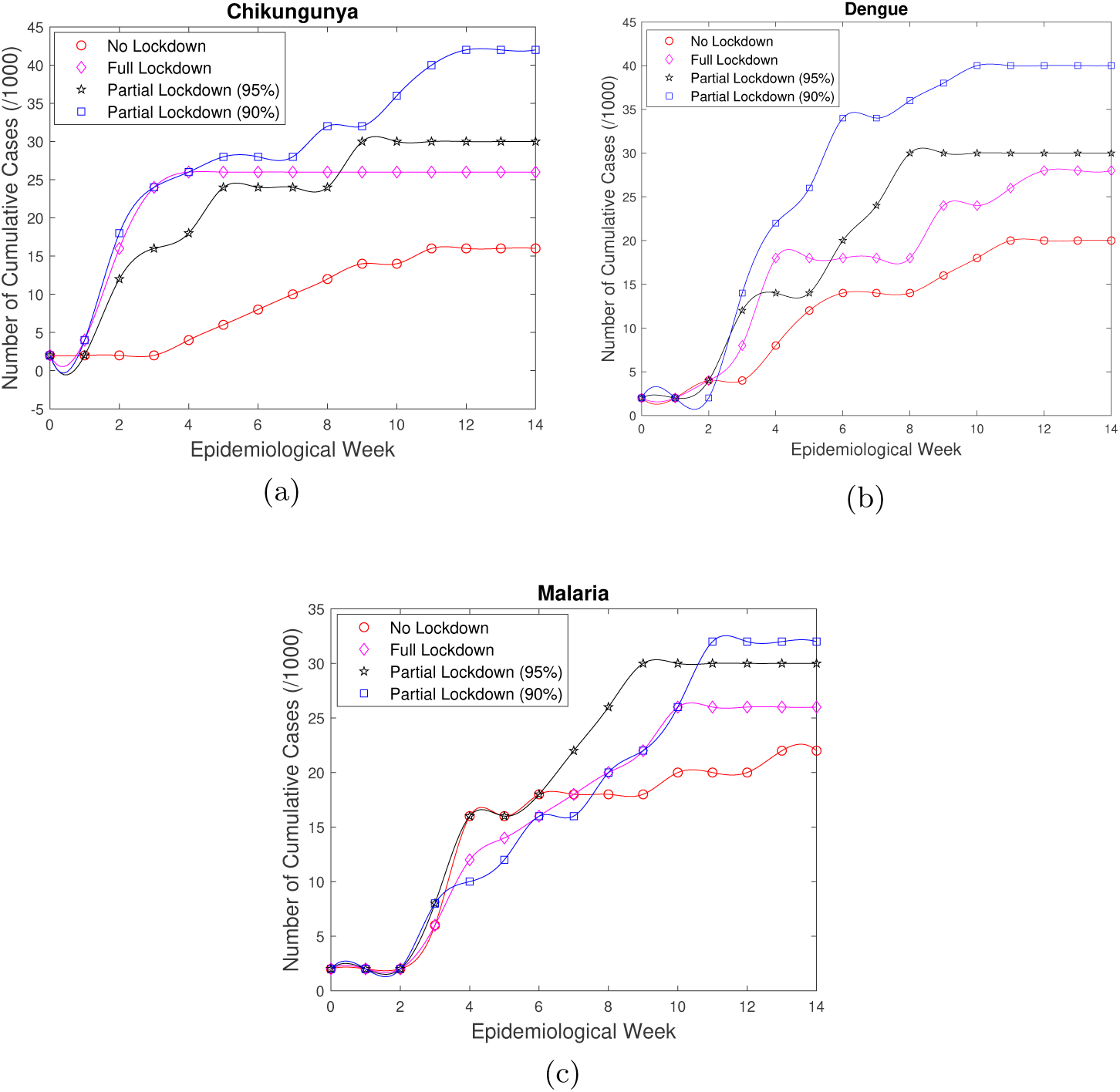
Epidemic progression of the three diseases in different lockdown scenarios.

#### Dengue

Dengue outbreaks have similar trajectories as chikungunya, except that in the no-lockdown condition, dengue spreads considerably faster than chikungunya (Figure 1b) though they share the same vector.

#### Malaria

The above results also apply to malaria but are less pronounced (Figure 1c).

### Decentralized intervention strategies are superior in lockdown

Intervention strategies for mosquito-borne diseases can be broadly classified into two categories: centralized and decentralized [9]. Centralized intervention strategies aim to reduce mosquito populations by directly attacking the mosquito breeding grounds. Decentralized intervention strategies work at an individual level where people are asked to adopt prophylactic measures like use of mosquito repellent creams, liquids, coils, mats, etc.

We tested the impact of Larval Source Management (LSM), a centralized strategy, against insecticide-treated nets (ITNs) + indoor residual spraying (IRS), a decentralized strategy for both full-lockdown and no-lockdown conditions. We report the results for chikungunya below.

Figure 2a shows that for no lockdown, LSM is more effective than ITN+IRS in mitigating the severity of the outbreak. Figure 2b indicates the reversal of this result in lockdown condition. Decentralized strategies prove to be more effective than centralized measures in case of a lockdown.

**Figure 2:**
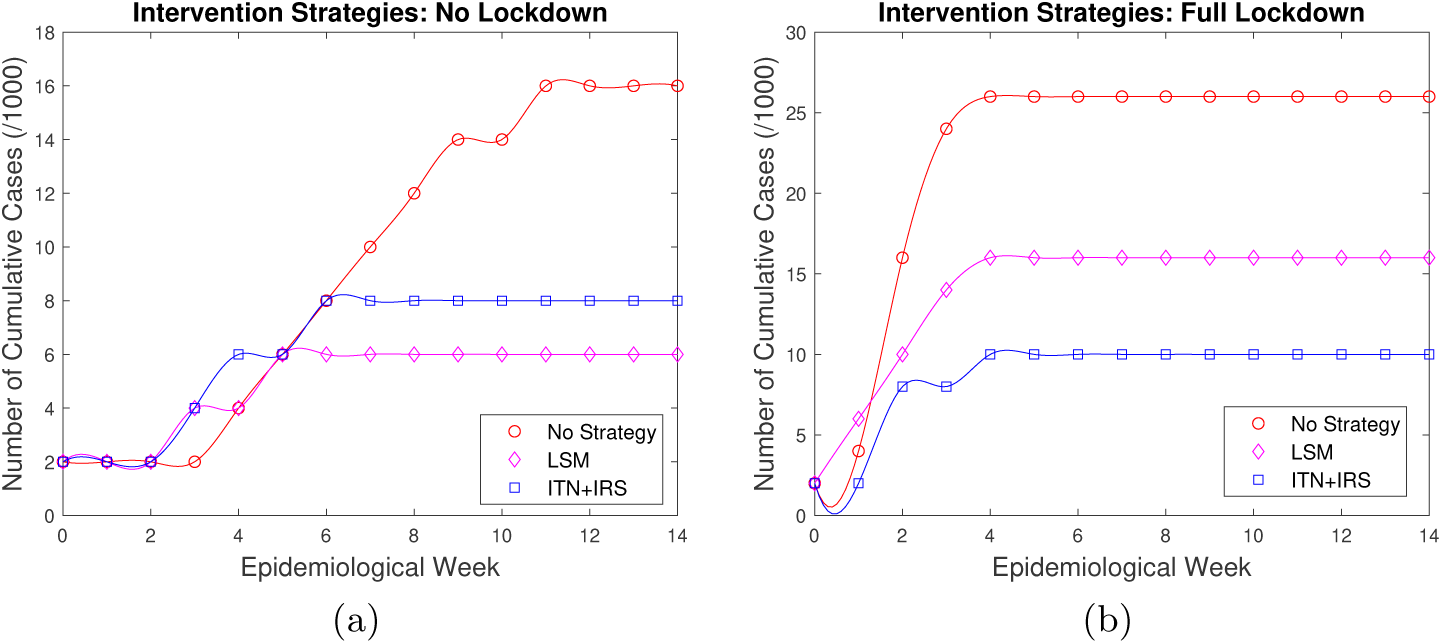
Prevention Strategies tested for chikungunya epidemic in lockdown vs no lockdown scenario.

## 3 Discussion

Mosquito-borne diseases continue to be a serious menace to humanity, and in fact are becoming more serious and widespread (seen in higher latitudes than formerly). Specifically, it is well known that perhaps 10% of all humans worldwide are infected with a mosquito-borne disease each year, and that as many as a million people die from it [5]. Thus, while humanity is in the midst of the COVID-19 pandemic and may even later find it prudent to enforce lockdowns to contain such epidemics, it is thus critical to understand how such lockdowns may effect the epidemiology of these diseases.

In the first part of this study, we investigated the impact of lockdowns on the epidemic trajectory of different mosquito-borne diseases by running simulations on our previously reported agent-based model [9]. We found that there is a consistent rise in the growth rates of these epidemics, in addition to a significant increase in the cumulative number of infected cases in lockdown scenarios. This drastic impact is a direct outcome of the increase in the probability of an infected mosquito biting multiple humans due to increased local population density and reduced human mobility on account of such lockdowns.

We also tested for partial lockdowns, analogous to real-world lockdowns which allow continued operations of essential and critical services, or lax observance of restrictions by some fraction of human society. We found the results to be much worse than with total lockdowns, as the total number of cases even doubled in some cases relative to the full-lockdown configuration. This increase in cases can be attributed to the increased mobility of infections over larger distances enabled by the few human agents still moving across the population in partial lockdowns, while also preserving the mosquitoes’ advantage in being able to infect more humans. This result suggests that additional care must be taken to see that essential workers who continue to be mobile in lockdowns do not carry mosquito-borne diseases, and that others who may carry such infections are not allowed to move around.

We further analyzed how the impact of lockdowns varies across three mosquito-borne diseases: chikungunya, dengue and malaria. We found that dengue outbreaks are more severe than chikungunya in any situation even though they share the same vector, *Aedes Aegypti*. This is because the dengue virus is transmissible from humans to mosquitoes even during late intrinsic incubation stage and early recovered stage. This in turn allows movement of infections by means of humans who are infectious but not showing symptoms (ironically in a manner similar to COVID-19). We also observed that the effects of lockdowns are less pronounced on malaria outbreaks, as they are spread by female *Anopheles* mosquitoes which are generally active during the night, a time where most humans are stationary with or without lockdowns.

Most mosquito-borne diseases are treatable but not easily curable, and still lack vaccines. It is widely accepted that the best strategy to control mosquito-borne epidemics is to reduce the mosquito population wholesale, or for humans to avoid mosquito bites at an individual level. Following this, we compared the effectiveness of two such strategies, LSM and ITNs+IRS, in reducing the number of infected cases. We discovered that while centralized strategies like LSM are more effective in no-lockdown scenarios, decentralized strategies such as ITNs+IRS are more effective in controlling outbreaks during lockdowns. We believe this is because decentralized measures directly address the reason behind the faster growth of epidemics in lockdowns by protecting the highly localised population density.

In accordance with this, it would appear prudent for policy-makers to guide the general public to be more proactive during lockdowns to take suitable personal measures to avoid mosquito bites.

We have not considered the reduced efficacy of the healthcare system on account of the preoccupation with COVID-19, or the general decline in health and immunity that may happen with reduced exercise and mobility under lockdown. We likewise do not consider the likely reduction in measures that are commonly taken to fight outbreaks of mosquito-borne diseases (such as cleaning and draining of stagnant bodies of water, and spraying of insecticides), though these are also likely. As such, it seems reasonable to expect that the risk and severity of outbreaks of mosquito-borne diseases will be even larger than suggested by our model.

The future course of the COVID-19 pandemic is still unclear at this time; while we may hope that it goes away very quickly, it may well sustain in some form for a long while to come, including by returning in a “second wave”, or by becoming a seasonal disease. Last but not least, it seems fair to surmise that even when COVID-19 is finally overcome, humanity will not have seen the last of such epidemics and pandemics that may entail the use of lockdowns. Thus, the issue of how to deal with mosquito-borne diseases while enforcing lockdowns will continue to be pertinent.

## 4 Methods

For this study, we have modified our previously reported agent-based model [9] to support multiple mosquito-borne diseases and to simulate social lockdowns. The previous model has been shown to outperform previous mathematical models, with the capability of predicting the trajectory of real mosquito-borne epidemics with 93% accuracy. The model has also proven its utility in estimating some hard-to-determine vector parameters such as mosquito sensory range and mortality rate, in addition to serving as a test bed for low-level policy designs such as intervention strategies for controlling epidemics and mosquito population.

We first briefly describe the design of our previous model, followed by the modifications we made to it for this study.

### Basic agent-based model

Our model works by capturing the complex relationship of human-infection-mosquito during an outbreak in urban environments by modelling them independently using simple adaptive rules. It is composed of three parts:

1. An infection life-cycle represented using a modified SEIR model with variable transition probabilities, capable of encapsulating a large proportion of mosquito-borne diseases.
2. Mosquito population distribution as a function of environmental factors such as temperature and precipitation. It explicitly models the evolution of every stage of the mosquito life-cycle (*eggs → larvae → pupae → mosquito*) to capture the complex vector dynamics.
3. Laws governing the movement of humans in an urban setting to capture the long-distance spatial movement of infection. Human agents are divided into four general categories based on their mobility patterns and frequency of exposure to mosquito breeding grounds.

The above three parts are then allowed to interact in a virtual environment which is built using real-world multi-layered GIS data, climate conditions and census information. The simulations run at the level of a small city, and agent granularity is at the level of an individual human and an individual mosquito. This lets us capture both the spatial and the temporal dynamics of such outbreaks. For more details on the model, we refer readers to our previous paper [9].

### Model modifications

We modified the model described above to study the progress of mosquito-borne diseases during lockdown scenarios. To accurately represent lockdowns as seen in our current circumstances, we tested for three different lockdown configurations:

1. *Full lockdown*: All human agents were made stationary and confined to their assigned buildings. In this situation, the mobility of infection is limited to mosquitoes.
2. *Partial lockdown* (*95%*): Similar to full lockdown, but 5% of the working human population is allowed movement to carry on with their regular movements. This represents the situation where lockdowns are accompanied by continued operation of essential services such as hospitals and government services, or lax observance of lockdowns by some people.
3. *Partial lockdown* (*90%*): Similar to the previous, but 10% of the working human population is allowed to carry on with their regular movements.

The model was previously tuned to only support *chikungunya* infection. We extended the model to support two additional prevalent mosquito-borne diseases: *dengue* and *malaria*. For modeling dengue and malaria epidemics, the parameters for infection and mosquito life-cycle, and infection transmission probabilities were adjusted accordingly. The altered parameters are summarized in Table 2. It should be noted that the dengue virus can also be passed from human to mosquito in its late intrinsic incubation stage, plus early recovery stage [11]. The existing SEIR model was modified to support this possibility. As seen in Table 2, the mosquito activity period also varies across diseases. This is because chikungunya and dengue share the same mosquito vector *Aedes Aegypti*, which usually operates during the day, in contrast to malaria, which is spread by night-biting *Anopheles* mosquitoes. We presume that 30% of the working human population works during the night instead of the day (as night-shift workers and the like).

**Table 2:** Modified ABM Parameters

| Parameter Description | Value<br>(Chikungunya) | Value<br>(Dengue) | Value<br>(Malaria) | References |
| --- | --- | --- | --- | --- |
| **Mosquito Parameters** |  |  |  |  |
| Active period | 7:00 a.m. to<br>6:00 p.m | 7:00 a.m. to<br>6:00 p.m | 6:00 p.m. to<br>7:00 a.m. | [12, 11, 3] |
| **Infection Transmission Cycle Parameters** |  |  |  |  |
| Transmission probability of<br>infection from human to mosquito | 0.275 | 0.275 | 0.3 | [1, 6] |
| **Infection Cycle Parameters:** |  |  |  |  |
| Number of days a human<br>spends in exposed state | 2 to 6 days | 4 to 10 days | 10 to 15 days | [12, 11, 2] |
| Number of days a human<br>spends in infected state | 4 to 7 days | 2 to 7 days | 5 to 7 days | [12, 11, 1, 2] |

We used the GIS and climate data of Saint Barthèlemy (a Caribbean island) to simulate a small tropical town. Finally, 20 rounds of simulations each were run for full-lockdown, partial-lockdown (95%), partial-lockdown (90%) and no-lockdown scenarios for each of the three diseases. Since the model has previously proven to be a reasonably effective representation of a real epidemic, we also used it to test and compare the efficacy of centralized and decentralized intervention strategies in controlling the chikungunya epidemic for full-lockdown and no-lockdown conditions.

### Statistical Analysis

Standard errors and confidence intervals for ABM simulations are poorly defined due to the failure of the *i*.*i*.*d*. assumption within a simulation run and the lack of concept of a “true” value for a virtual population. As such, all the results reported in the study are within the 95% range interval (i.e., the interval wherein 95% of simulation results are contained) as suggested by [4].

## Data Availability

All data required are available as noted in the paper.

https://github.com/akshayjin/chikungunya_simulation

## Code and Data Availability

The ABM simulations are implemented on GAMA platform v1.6 [10]. The modified model source code is available on our GitHub repository [8]. The authors declare that all other data supporting the findings of this study are available within the article and its Supplementary Information files, or are available from the authors upon request.

## Acknowledgements

The work of the second author was supported in part by an AWS Machine Learning Research Award from Amazon.

## Author Contributions

A.J. programmed and performed the agent-based simulations. S.R. conceptualised the problem and wrote the first draft. Both authors wrote the manuscript.

## Competing Interests

The authors declare that they have no competing interests.

## Notes

### Competing Interest Statement

The authors have declared no competing interest.

## References

[1] Bershteyn, A., Gerardin, J., Bridenbecker, D., Lorton, C. W., Bloedow, J., Baker, R. S., Chabot-Couture, G., Chen, Y., Fischle, T., Frey, K., and et al. Implementation and applications of emod, an individual-based multi-disease modeling platform. Pathogens and Disease 76, 5 (Jan 2018). Available at https://idmod.org/docs/malaria/parameter-configuration.html.

[2] Bleijs, D. Aedes albopictus. http://www.chikungunyavirusnet.com/aedes-albopictus.html, 2014. Accessed: 2016-04-12.

[3] Bleijs, D. Aedes aegypti. http://www.denguevirusnet.com/aedes-aegypti.html, 2016. Accessed: 2016-04-12.

[4] Bobashev, G. V., and Morris, R. J. Uncertainty and inference in agent-based models. In 2010 Second International Conference on Advances in System Simulation (2010), IEEE, pp. 67–71.

[5] Caraballo, H., and King, K. Emergency department management of mosquito-borne illness: malaria, dengue, and west nile virus. Emergency medicine practice 16, 5 (2014), 1–23.

[6] Dommar, C. J., Lowe, R., Robinson, M., and Rodó, X. An agent-based model driven by tropical rainfall to understand the spatiotemporal heterogeneity of a chikungunya outbreak. Acta Tropica 129 (Jan. 2014), 61–73.

[7] Green, M. S., Swartz, T., Mayshar, E., Lev, B., Leventhal, A., Slater, P. E., and Shemer, J. When is an epidemic an epidemic? The Israel Medical Association journal: IMAJ 4, 1 (2002), 3–6.

[8] Jindal, A. Chikununya Simulation. https://github.com/akshayjin/chikungunya_simulation, Mar 2020. Accessed: 2020-03-27.

[9] Jindal, A., and Rao, S. Agent-based modeling and simulation of mosquito-borne disease transmission. In Sixteenth International Conference on Autonomous Agents and Multiagent Systems (AAMAS 2017) (São Paulo, Brazil, May 2017), pp. 426–435. http://www.ifaamas.org/Proceedings/aamas2017/pdfs/p426.pdf.

[10] Taillandier, P., Gaudou, B., Grignard, A., Huynh, Q.-N., Marilleau, N., Caillou, P., Philippon, D., and Drogoul, A. Building, composing and experimenting complex spatial models with the gama platform. GeoInformatica 23, 2 (2019), 299–322.

[11] WHO. Dengue and severe dengue. https://www.who.int/news-room/fact-sheets/detail/dengue-and-severe-dengue, Mar 2020. Accessed: 2020-03-27.

[12] WHO. Fact sheet about Malaria. https://www.who.int/news-room/fact-sheets/detail/malaria, Jan 2020. Accessed: 2020-03-27.

